# Correlates of and changes in aerobic physical activity and strength training before and after the onset of COVID-19 pandemic in the UK – findings from the HEBECO study

**DOI:** 10.1101/2021.01.16.21249925

**Authors:** Aleksandra Herbec, Verena Schneider, Abi Fisher, Dimitra Kale, Lion Shahab, Phillippa Lally

**Affiliations:** Department of Behavioural Science and Health, University College London, UK; Clinical, Educational and Health Psychology, University College London, UK

**Keywords:** aerobic activity, strength training, covid-19, UK, cross-sectional, changes, MSA, MVPA

## Abstract

**Objectives:** Understanding changes in moderate to vigorous aerobic physical activity (MVPA) and strength training (MSA) from before to after (pre-/post-) the onset of the Covid-19 pandemic in the UK (first lockdown) and their correlates can inform interventions.

**Methods:** Cross-sectional analysis of retrospective and concurrent data on MVPA/MSA pre- and post-Covid-19 (until 14^th^ June 2020) among 2,657 UK adults. The associations between socio-demographic and health characteristics, MVPA/MSA pre-Covid-19, living and exercise conditions and meeting WHO recommended levels for MVPA/MSA/both (vs meeting neither), and changes in MVPA/MSA from pre-to post-Covid-19 following stratification for pre-Covid-19 MVPA/MSA levels were evaluated.

**Results:** A third of adults maintained (30.4%), decreased (36.2%) or increased (33.4%) their MVPA levels post-Covid-19. For MSA, the percentages were 61.6%, 18.2%, and 20.2%, respectively. MVPA increased or decreased by an average of 150min/week, and MSA by 2 days/week. Meeting both MSA+MVPA recommendations during lockdown (vs. meeting neither) was positively associated with meeting MVPA+MSA pre-lockdown (aOR=16.11,95%CI=11.24-23.07), and post-16-years of age education (aOR=1.57,1.14-2.17), and negatively associated with being obese (aOR=0.49,0.33-0.73), older age (65+ vs ≤34; aOR=.53,.32-.87), and annual household income <50.000GBP (vs ≥50.000GBP; aOR=0.65,0.46-0.91). The odds for decreasing MVPA were significantly lower for white ethnicity, post-16-years of age education, access to garden/balcony, and higher for those who were in total isolation. The odds for decreasing MSA were significantly higher for those who were overweight or obese.

**Conclusion:** Aerobic and strength training were differently impacted during the first UK lockdown, with poorer outcomes associated with older age, lower education, and higher body mass index.

What are the new findings

- At the start of the Covid-19 pandemic, a third of UK adults decreased and a third increased moderate-to-vigorous physical activity; the corresponding proportions for muscle strength training were 18% and 20%.
- Older adults were more likely to maintain inactivity from before to start of the pandemic.
- Adults with higher body mass index were at risk of not meeting WHO recommendations for weekly levels of strength and aerobic training, and at decreasing muscle strength training in this period.
- Those with lower incomes, no post-16-years of age education, ethnic minority background and living in total isolation had poorer physical activity outcomes.

## Introduction

Physical activity has been negatively affected by the Covid-19 pandemic, which could have major implications for general health and Covid-19 outcomes^1,2,3^,. As the Covid-19 pandemic continues and new lockdowns are introduced it is paramount to understand how aerobic and muscle-strength training were affected during the pandemic so far, and which group in the society may require most support.

Adults who engage in less than 30min of moderate physical activity per week are considered to be inactive ^4,5^.For best health outcomes adults are recommend to engage in moderate-to-vigorous intensity aerobic physical activity (MVPA; i.e. activities that increase heart rate and make one feel warmer) for at least 150 minutes/week, as well as in muscle-strengthening activity (MSA; e.g., strength/resistance training) for at least two days/week^6^. MVPA and MSA both independently and combined lower morbidity and mortality ^7-11,12^.

The Covid-19 pandemic and the social distancing measures introduced in the UK on 23^rd^ March 2020 have affected many opportunities to engage in physical activity ^13^. Many sports centres (e.g. gyms) were temporarily closed and team sports banned, while access to outdoors spaces (e.g. remote national parks) was greatly reduced. On the other hand, some lockdown measures could facilitate exercising, including the introduction of remote working. Furthermore, exercising outdoors was listed as one of the few activities that were still permitted even during the strictest lockdown in the UK, alongside shopping for essential items or medicine and going to work.

Physical activity during the Covid-19 pandemic declined^14^, but a small minority of adults increased their activity^15^. Female gender, lower income, older age, health conditions, perceived risks and poorer mental health were associated with lower physical activity levels^15-18^. However, research to date has focused primarily on MVPA or failed to distinguish between MVPA or MSA^15,19,20^and few studies accounted for confounders that could be relevant during the pandemic, such as having access to indoor and outdoor exercising space or the household makeup^15^.

### Aims

We characterised MVPA and MSA levels during the periods *before* and *after* (pre- /post-) the Covid-19 pandemic had started in the UK, with the latter covering the period of the first strictest UK lockdown. The research questions (RQs) were:

- RQ1: What were the levels and changes in MVPA and MSA among UK adults from pre- to post-Covid-19?
- What socio-demographic, environmental and health factors were associated with: RQ2: meeting WHO recommendations for MVPA and MSA post-Covid-19? RQ3: (i) decrease in MVPA among those who were active, and (ii) increase in MVPA among those who were inactive from pre- to post-Covid-19? RQ4: (i) decrease in MSA among those who were active, and (ii) increase in MSA among those who were inactive, from pre- to post-Covid-19?

## Methods

### Study Design

The study involved cross-sectional data analysis from the baseline survey of the HEalth BEhaviours during the COvid-19 pandemic (HEBECO) study (https://osf.io/sbgru/). Data were collected using REDCap at UCL (Harris et al, 2019). The study protocol was preregistered (https://osf.io/ejghs/). Departures from the protocol are explained in Supplementary Materials 1 (https://osf.io/w7vbf/). The results from all the pre-registered analyses not reported here are available on Open Scinece Framework: https://osf.io/2ujxq/. The reporting follows the STROBE guidelines.^21^

### Participants

Recruitment involved a UK-wide campaign including paid and unpaid posts on social media and information shared through the networks of Cancer Research UK, Public Health England, other charities, sports clubs, universities, and local authorities. Participants with complete data who enrolled into the baseline between 5^th^ May 2020 (start of the data collection for all study variables) until 14^th^ June 2020 (prior to easing of restrictions of the first UK lockdown) were included.

### Measures

For the wording of all measures see https://osf.io/bja7g/ and Supplementary Materials 2. At baseline participants indicated the time when their lives started to be affected by the Covid-19 pandemic in any way, which was used as an anchor to assess before/since (pre-/post-Covid-19) MVPA and MSA. Almost half (44.0%, weighted) of adults selected the second half of March, followed by 26.9% who selected the first half of March.

#### Meeting WHO recommendations for MVPA and MSA

The pre-/post-Covid-19 MVPA and MSA were measured using the same set of questions^12,22.^ For MVPA, participants were first asked “In the month Before/Since Covid-19, on average, HOW MANY TIMES PER WEEK did you do at least 15 MINUTES or more of moderate or vigorous AEROBIC PHYSICAL ACTIVITY? Examples: brisk walk, jogging, dancing, cycling for recreation or commute, swimming, team or racket sports.” The answers were capped at 14+, equivalent to at least twice per day. The 15 minute period was selected as it corresponds to the minimum amount of PA needed for mortality reduction and extending lifespan^23^.Those who engaged in at least one such session were asked “In the month BEFORE/Since COVID-19, HOW LONG (in MINUTES) was your average session of moderate or vigorous AEROBIC PHYSICAL ACTIVITY? Do NOT include strength training.” Total weekly MVPA was computed by multiplying the session number by the minutes.

MSA activity was assessed by a single question: “Before/Since Covid-19, on average, on HOW MANY DAYS PER WEEK did you do STRENGTH TRAINING? Examples: Pilates, push-ups, squats, yoga, and exercises involving free weights, weight machines or elastic band.” The answers were capped at 4+ given that the recommendations were for 2 days of MSA per week.

Data on MVPA and MSA were used to categorise participants, separately for pre-/post-Covid-19, into four groups based on whether they met the WHO recommendations for MSA (at least 2 days/week) and MVPA (≥150 minutes/week)^6^ : meeting neither, meeting MSA only, meeting MVPA only, or meeting recommendations for both MSA and MVPA.

#### Change in MVPA and MSA

Changes in MVPA (RQ 3) and MSA (RQ 4) were calculated by subtracting the pre-Covid-19 levels of MSA or MVPA from the post-Covid-19 levels. Participants were categorised into: maintenance (change in MVPA <20 min; change in MSA=0), decrease (MVPA by ≥20min; MSA by ≥1 day) or increase in activity (MVPA by ≥20min; MSA by ≥1 day). Dichotomous variables were then creased: decrease (vs maintain/increase) and increase (vs maintain/decrease) in MVPA and MSA.

To minimise the bias due to ceiling and floor effects when assessing changes to MVPA and MSA, the analyses for RQs 3 and 4 were conducted after stratifying for pre-Covid-19 MVPA and MSA. Participants were categorised into: inactive MVPA (<30min of MVPA/week^4,5^) or MSA (0 days of MSA/week) and active (≥30 min/week of MVPA; and ≥ 1 day/week of MSA) pre-Covid-19.

Participants who were active post-Covid-19 were also asked “Do you do the same FORM of exercise as you did before the COVID-19? (even if it is in a different location)?” The answer options were: None of the same/some of the same/about half the same/mostly the same/exactly the same.

#### Explanatory factors and correlates

Socio-demographic characteristics and living conditions assessed were: gender (female/other), ethnicity (white/non-white); education (post-16-years of age/other); employed (yes/no); furloughed/laid off (yes/no); income (low-middle <50 000 GBP/high ≥50 000 GBP/prefer-not-to-say); age (in decades was entered as a continuous variable where it met assumptions, or as a 3-level categorical variable: ≤34, 35-64, 65+).

We assessed health behaviours, health and living conditions that could impact on MVPA or MSA levels: self-reports of any condition that limited physical activity (yes/no); body mass index (BMI^22^: normal and underweight ≤24.99/overweight 25-29.99/ obese ≥30); smoking status (current smoker vs not)^24^; weekly frequency of alcohol drinking^25^; deterioration in psychological wellbeing from pre- to post-Covid-19 (yes/no); isolation status (total isolation/other); participants’ perceived risk of Covid-19 to their health (‘no or minor risk’/other); access to a garden or balcony big enough to exercise comfortably (yes/no); access to a public park/green space that is within a walking distance and open during Covid-19 (yes/no); living with children aged ≤15 (yes/no); living with vulnerable persons (persons over the age of 70, in poor health, or who may be vulnerable to Covid-19; yes/not).

Model included two time co-variates to account for weather changes: enrolment time (up until 15th May, the second half of May, the first half June), and time when Covid-19 started to affect individuals (before mid-March/later).

### Analysis

Analyses were conducted in SPSS 26 with the data weighted using the 2018 Census and APS mid-year estimates for age, gender, ethnicity, country of living and household income. The analysis used weights trimmed to top 98^th^ percentile to minimise the impact of extremely high weights ^26^. Differences between the included and excluded participants were assessed with chi-squared for categorical and t-tests for continuous data.

For RQ1, descriptive statistics were computed to characterise the levels of MVPA, MSA, and inactivity pre-/post-Covid-19 and changes in exercise form. For RQ2 univariate and fully adjusted multinomial regression models were computed. For RQ3 and RQ4 the sample was stratified using pre-Covid-19 MVPA or MSA levels to assess outcomes of interest using univariate and adjusted logistic regression models. Sensitivity analyses involved replicating the analyses using unweighted data to check for the robustness of the results, and for RQ3 using differ cut-off values of 15mins and 30mins.

Family-wise error was corrected for by using the Benjamini-Hochberg procedure separately for each research question (Benjamini and Hochberg, 1995).

## Results

The final analytic sample included 2,657 adults with complete study data (weighted N=2,442). Table 1 present comparisons of the excluded and included sample. The included weighted sample comprised 52% females, 90.5% of white ethnicity, and 67.0% with high school education or higher.

**Table 1.**
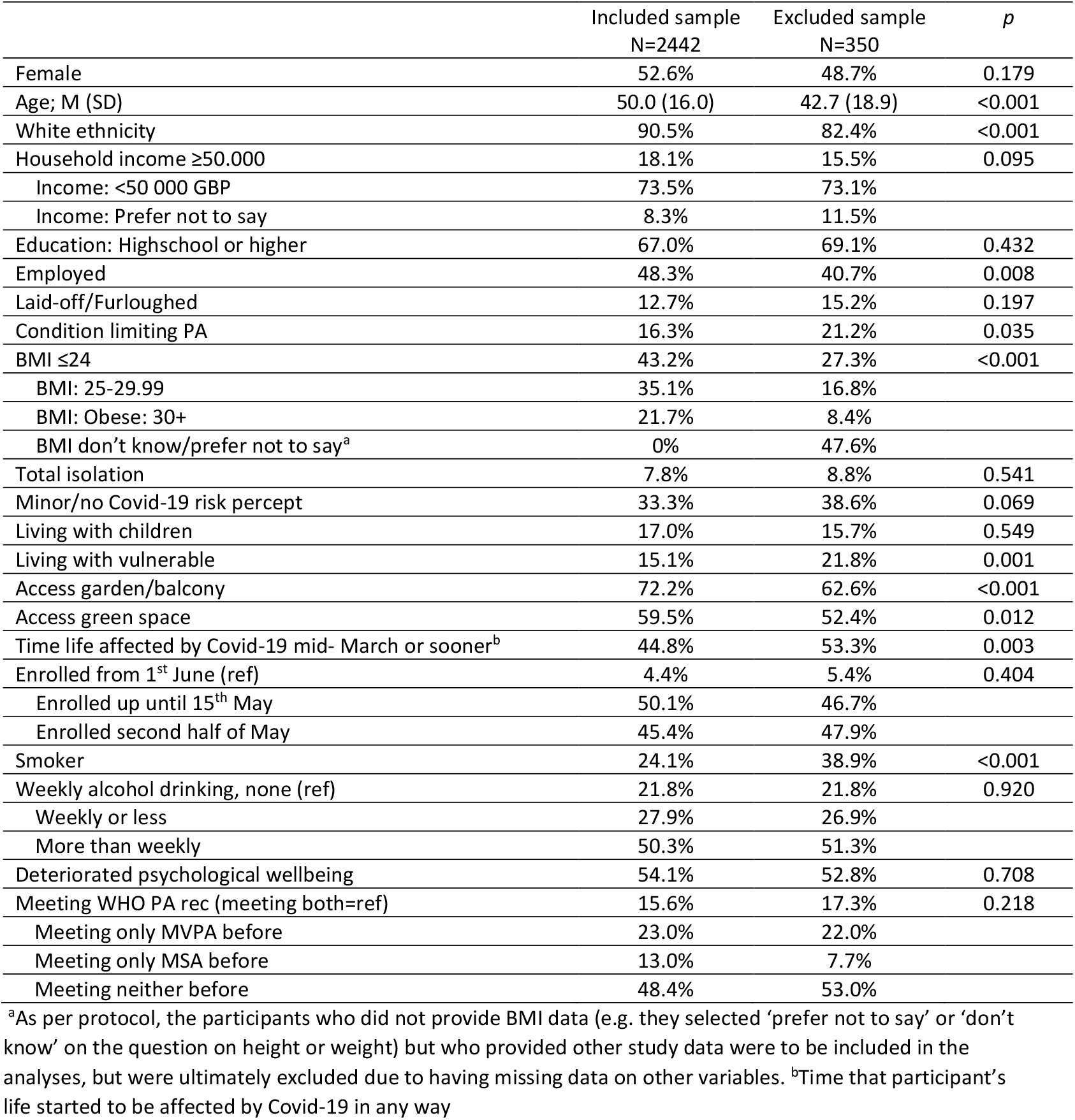
Sample characteristics (weighted).

|  | Included sample<br>N=2442 | Excluded sample<br>N=350 | *p* |
| --- | --- | --- | --- |
| Female | 52.6% | 48.7% | 0.179 |
| Age; M (SD) | 50.0 (16.0) | 42.7 (18.9) | <0.001 |
| White ethnicity | 90.5% | 82.4% | <0.001 |
| Household income ≥50,000 | 18.1% | 15.5% | 0.095 |
| Income: <50,000 GBP | 73.5% | 73.1% |  |
| Income: Prefer not to say | 8.3% | 11.5% |  |
| Education: Highschool or higher | 67.0% | 69.1% | 0.432 |
| Employed | 48.3% | 40.7% | 0.008 |
| Laid-off/Furloughed | 12.7% | 15.2% | 0.197 |
| Condition limiting PA | 16.3% | 21.2% | 0.035 |
| BMI ≤24 | 43.2% | 27.3% | <0.001 |
| BMI: 25-29.99 | 35.1% | 16.8% |  |
| BMI: Obese: 30+ | 21.7% | 8.4% |  |
| BMI don't know/prefer not to say<sup>a</sup> | 0% | 47.6% |  |
| Total isolation | 7.8% | 8.8% | 0.541 |
| Minor/no Covid-19 risk percept | 33.3% | 38.6% | 0.069 |
| Living with children | 17.0% | 15.7% | 0.549 |
| Living with vulnerable | 15.1% | 21.8% | 0.001 |
| Access garden/balcony | 72.2% | 62.6% | <0.001 |
| Access green space | 59.5% | 52.4% | 0.012 |
| Time life affected by Covid-19 mid- March or sooner<sup>b</sup> | 44.8% | 53.3% | 0.003 |
| Enrolled from 1<sup>st</sup> June (ref) | 4.4% | 5.4% | 0.404 |
| Enrolled up until 15<sup>th</sup> May | 50.1% | 46.7% |  |
| Enrolled second half of May | 45.4% | 47.9% |  |
| Smoker | 24.1% | 38.9% | <0.001 |
| Weekly alcohol drinking, none (ref) | 21.8% | 21.8% | 0.920 |
| Weekly or less | 27.9% | 26.9% |  |
| More than weekly | 50.3% | 51.3% |  |
| Deteriorated psychological wellbeing | 54.1% | 52.8% | 0.708 |
| Meeting WHO PA rec (meeting both=ref) | 15.6% | 17.3% | 0.218 |
| Meeting only MVPA before | 23.0% | 22.0% |  |
| Meeting only MSA before | 13.0% | 7.7% |  |
| Meeting neither before | 48.4% | 53.0% |  |

<sup>a</sup>As per protocol, the participants who did not provide BMI data (e.g. they selected 'prefer not to say' or 'don't know' on the question on height or weight) but who provided other study data were to be included in the analyses, but were ultimately excluded due to having missing data on other variables. <sup>b</sup>Time that participant's life started to be affected by Covid-19 in any way

Below are results for the weighted fully adjusted analyses. Results from univariable analyses are reported in Supplementary Materials 3, and from sensitivity analyses in Supplementary Materials 4 and 5, and have not changed the conclusions.

### Research Question 1: Changes in MVPA and MSA pre- to post-Covid-19

Table 2 presents data on physical activity levels before and since Covid-19. Pre-Covid-19 17.6% of adults engaged in no MVPA and MSA, 55.3% engaged in no MSA, and 19.4% engaged in no MVPA. Just under 15% of adults met the recommended levels of both MVPA and MSA pre- and post- Covid-19. The proportion of those who had no MVPA or MSA activity increased minimally (17.4% to 22.1%) which was primarily driven by declines in MVPA. Among those who were active post-Covid-19, 41.5% continued with the mostly or exactly activity form as pre-Covid-19.

**Table 2.** Aerobic physical activity (MVPA) and strength training (MSA) before and since COVID-19.

|  | Before Covid-19<sup>1</sup> | Since Covid-19<sup>1</sup> |
| --- | --- | --- |
| **No activity (0 sessions MVPA and 0 days MSA) % (N)** | 17.6 (430) | 22.1 (540) |
| **MVPA** |  |  |
| MVPA 0 sessions % (N) | 19.4 (473) | 24.8 (604) |
| MVPA min/week among active; Median (IQR) | 125 (210.0) | 150 (220.0) |
| Mean (SD) | 238.1 (424.0) | 230.9 (398.8) |
| MVPA min/week among entire sample; Median (IQR) | 90 (180.0) | 90 (224.5) |
| Mean (SD) | 196.2 (395.4) | 179.8 (364.7) |
| **MSA** |  |  |
| MSA % (N) |  |  |
| 0 das/week | 55.3 (1557) | 56.2 (1584) |
| 1 day/week | 11.1 (314) | 9.6 (271) |
| 2 days/week | 10.4 (293) | 7.9 (223) |
| 3 days/week | 9.0 (253) | 7.6 (213) |
| 4+ days/week | 7.0 (197) | 11.3 (320) |
| MSA Median (IQR) | 0 (2) | 0 (2) |
| **Meet guidelines % (N)** |  |  |
| Neither MVPA nor MSA | 45.1 (1271) | 44.6 (1256) |
| Meet MVPA ($\geq 150$min/week) only | 21.3 (599) | 21.2 (598) |
| Meet MSA ($\geq 2$ days/week) only | 11.7 (330) | 11.9 (336) |
| Meets both MVPA and MSA | 14.5 (410) | 14.8 (416) |
| **Change in form of exercise among those who exercise % (N)** |  |  |
| None of the same | - | 17.1 (331) |
| Some of the same | - | 32.7 (632) |
| About half the same | - | 8.7 (168) |
| Mostly the same | - | 26.0 (503) |
| Exactly the same | - | 15.5 (299) |

From before to since Covid-19 similar proportions of adults maintained (30.4%), decreased (36.2%) or increased (33.5%) their weekly MVPA. Among those who were active pre-Covid-19, 46.7% decreased MVPA. The decrease was of on average 219min/week. Among those who were inactive pre-Covid-19, 29.7% increased MVPA, with an average increase of 144min/week.

Maintenance of MSA was relatively more common (61.6%), with 18.2% decreasing and 20.2% increasing the number of days they engaged in MSA. Among those who were active pre-Covid-19 (40.2%), 45.1% decreased MSA, with an average decrease of 1.9 days/week. Among those who engaged in no MSA pre-Covid-19 (59.8%), 16.4% increased, with an average increase of 2.5 days/week.

### Research Question 2: Predictors of meeting WHO recommendations

Table 3 presents results from fully adjusted models. Being aged 65+, having a lower pre-Covid-19 household income, having a condition limiting physical activity, being obese, living in total isolation and deterioration in psychological wellbeing were associated with lower odds, and having at least high school education or meeting the WHO recommended levels of MVPA, MSA or both pre-Covid-19 was associated with greater odds of meeting both MVPA and MSA WHO guidelines since Covid-19 compared with not meeting either.

**Table 3.** Predictors of meeting both, MVPA only and MSA only WHO guidelines since Covid-19 in comparison to not meeting WHO guidelines for MVPA and MSA (reference, n=1178). Results from fully adjusted models on weighted data with BH FDR correction of p-values (significant in bold).

|  | Both MVPA and MSA (n=388) |  |  |  | MVPA only (n=570) |  |  |  | MSA only (n=306) |  |  |  |
| --- | --- | --- | --- | --- | --- | --- | --- | --- | --- | --- | --- | --- |
|  | aOR | 95% CI |  | p | aOR | 95% CI |  | p | aOR | 95% CI |  | p |
| Female | 1.08 | 0.82 | 1.41 | 0.588 | 0.79 | 0.62 | 1.00 | 0.049 | 1.02 | 0.77 | 1.35 | 0.906 |
| Age ≤34 |  |  |  |  |  |  |  |  |  |  |  |  |
| Age: 35-64 | 0.70 | 0.49 | 0.99 | 0.047 | 1.11 | 0.78 | 1.56 | 0.566 | **0.56** | **0.38** | **0.82** | **0.003** |
| Age: 65+ | **0.53** | **0.32** | **0.87** | **0.012** | 1.10 | 0.71 | 1.70 | 0.676 | 0.99 | 0.60 | 1.61 | 0.961 |
| White ethnicity | 1.61 | 1.03 | 2.52 | 0.036 | 1.57 | 0.99 | 2.49 | 0.054 | 0.72 | 0.47 | 1.10 | 0.130 |
| Household income ≥50.000 |  |  |  |  |  |  |  |  |  |  |  |  |
| Income: <50 000 GBP | **0.65** | **0.46** | **0.91** | **0.011** | 0.71 | 0.52 | 0.96 | 0.028 | 1.13 | 0.76 | 1.67 | 0.556 |
| Income: Prefer not to say | **0.44** | **0.25** | **0.77** | **0.004** | 0.63 | 0.38 | 1.03 | 0.065 | 0.88 | 0.49 | 1.59 | 0.666 |
| Education: Highschool or higher | **1.57** | **1.14** | **2.17** | **0.006** | 1.04 | 0.81 | 1.35 | 0.745 | **1.74** | **1.24** | **2.45** | **0.002** |
| Employed | 0.76 | 0.55 | 1.05 | 0.094 | 1.09 | 0.82 | 1.43 | 0.562 | 1.06 | 0.75 | 1.49 | 0.734 |
| Laid-off/Furloughed | 0.79 | 0.51 | 1.23 | 0.295 | 1.05 | 0.71 | 1.54 | 0.813 | 1.14 | 0.71 | 1.83 | 0.584 |
| Condition limiting physical activity | **0.44** | **0.28** | **0.71** | **0.001** | **0.43** | **0.29** | **0.63** | **<0.001** | 0.97 | 0.65 | 1.45 | 0.881 |
| BMI ≤24 |  |  |  |  |  |  |  |  |  |  |  |  |
| BMI: 25-29.99 | 0.81 | 0.60 | 1.10 | 0.176 | 1.01 | 0.78 | 1.32 | 0.926 | 0.72 | 0.53 | 0.99 | 0.043 |
| BMI: Obese: 30+ | **0.49** | **0.33** | **0.73** | **<0.001** | 0.87 | 0.64 | 1.19 | 0.394 | **0.37** | **0.24** | **0.56** | **<0.001** |
| Total isolation | **0.44** | **0.23** | **0.83** | **0.011** | **0.26** | **0.14** | **0.50** | **<0.001** | 0.83 | 0.49 | 1.40 | 0.478 |
| Minor/no Covid-19 risk percept | 1.00 | 0.74 | 1.34 | 0.977 | 1.20 | 0.93 | 1.55 | 0.167 | 0.94 | 0.68 | 1.30 | 0.718 |
| Living with children | 0.81 | 0.57 | 1.16 | 0.254 | 0.75 | 0.55 | 1.03 | 0.079 | 0.66 | 0.44 | 0.99 | 0.045 |
| Living with vulnerable | 1.17 | 0.82 | 1.68 | 0.395 | 0.86 | 0.61 | 1.20 | 0.366 | 0.87 | 0.58 | 1.30 | 0.499 |
| Access garden/balcony to exercise comfortably in | 1.10 | 0.81 | 1.51 | 0.533 | 1.02 | 0.79 | 1.33 | 0.861 | 0.80 | 0.58 | 1.09 | 0.159 |
| Access green space within walking distance | 1.27 | 0.96 | 1.69 | 0.096 | 1.31 | 1.03 | 1.66 | 0.028 | 0.91 | 0.68 | 1.23 | 0.548 |
| Smoker | 0.72 | 0.51 | 1.02 | 0.062 | 0.93 | 0.70 | 1.22 | 0.593 | 0.80 | 0.57 | 1.14 | 0.218 |
| Weekly alcohol drinking, none (ref) |  |  |  |  |  |  |  |  |  |  |  |  |
| Weekly or less | 1.00 | 0.68 | 1.49 | 0.988 | 1.43 | 1.02 | 2.00 | 0.038 | 1.06 | 0.71 | 1.59 | 0.782 |
| More than weekly | 1.04 | 0.73 | 1.50 | 0.811 | 1.28 | 0.94 | 1.74 | 0.121 | 0.89 | 0.61 | 1.29 | 0.528 |
| Deteriorated psychological wellbeing | **0.56** | **0.43** | **0.73** | **<0.001** | **0.71** | **0.56** | **0.89** | **0.003** | 0.72 | 0.55 | 0.96 | 0.025 |
| Meeting WHO PA<sup>a</sup> rec pre-Covid (meeting neither=ref) |  |  |  |  |  |  |  |  |  |  |  |  |
| Meeting both before | **16.11** | **11.24** | **23.07** | **<0.001** | **2.31** | **1.58** | **3.39** | **<0.001** | **3.72** | **2.45** | **5.62** | **<0.001** |
| Meeting only MVPA before | **3.88** | **2.64** | **5.70** | **<0.001** | **7.57** | **5.82** | **9.84** | **<0.001** | 1.47 | 0.94 | 2.30 | 0.092 |
| Meeting only MSA before | **6.38** | **4.26** | **9.54** | **<0.001** | 0.74 | 0.44 | 1.25 | 0.264 | **9.61** | **6.70** | **13.77** | **<0.001** |

a=meeting World Health Physical activity recommendations before the first Covid-19 lockdown in the UK. Notes: All models were also adjusted for the time of enrolment and time when Covid-19 started to affect individuals in any way. Following the BH correction the highest p-value that met the threshold for significance was p=0.019

Having a condition that limited physical activity, being in total isolation and deterioration in psychological wellbeing was associated with lower odds, and meeting the WHO recommended levels of MVPA or both MVPA and MSA was associated with greater odds of meeting MVPA only post-Covid-19. Being aged 35-64, and having obesity (BMI≥30) was associated with lower odds, while higher education, and meeting both MVPA and MSA or only MSA recommendations pre-Covid-19 was associated with meeting the recommendations for MSA only post-Covid-19.

### RQ 3: Associations with changes in MVPA

Older, white participants with at least high school education and access to a garden/balcony to exercise comfortably in were less likely to decrease MVPA activity (Table 4). Those with conditions limiting PA, living in total isolation and experiencing a deterioration in psychological wellbeing during lockdown were more likely to decrease MVPA activity from pre-Covid levels.

**Table 4.**
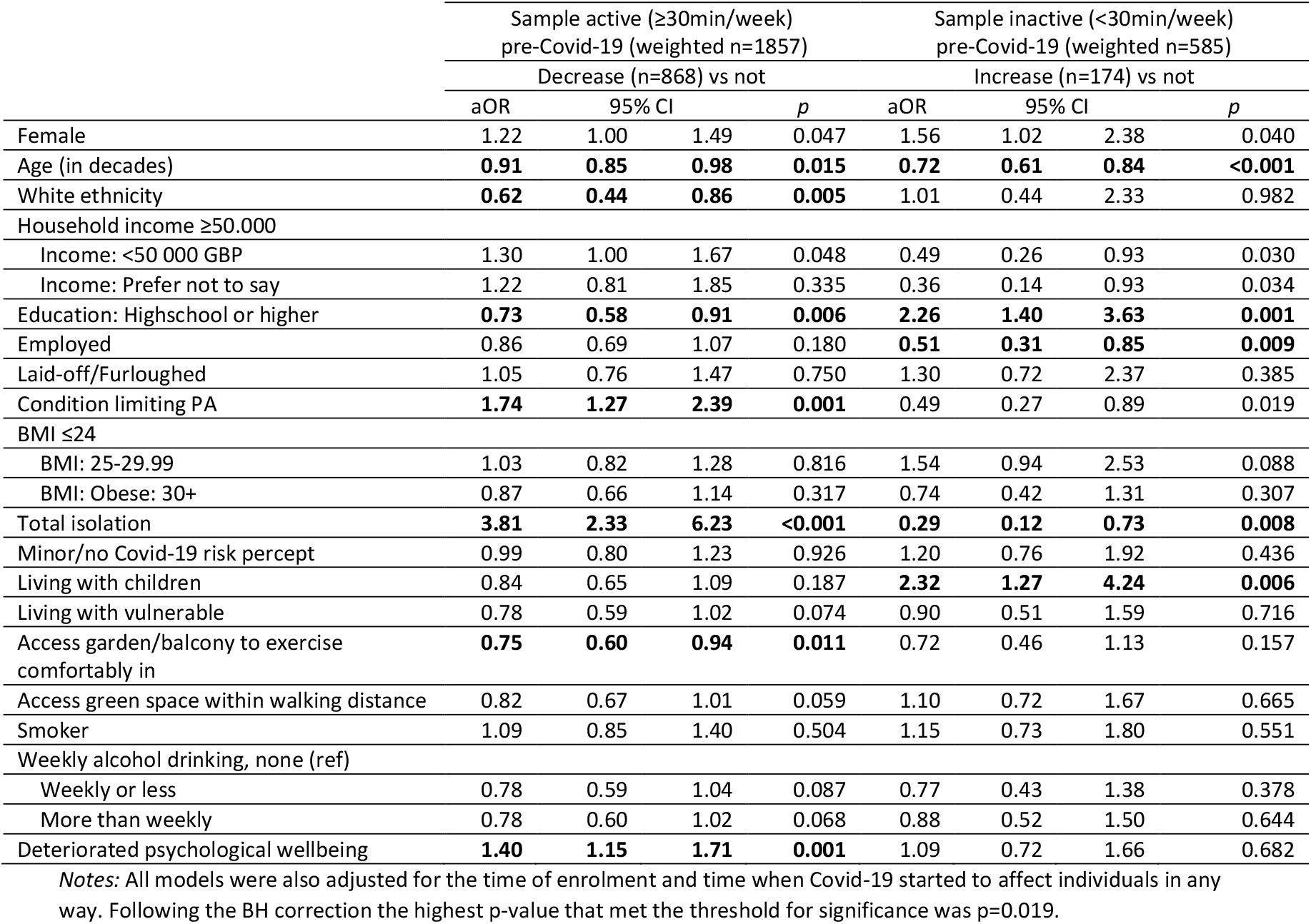
Independent associations of change in MVPA. Segmented analyses of active sample (pre-Covid-19 MVPA activity ≥30min; predicting decrease by ≥20min) and inactive sample (pre-Covid-19 MVPA activity <30min; predicting increase by ≥20min).

White ethnicity, being employed during Covid-19, and living in total isolation were associated with lower odds of increasing MVPA among this group. At least high school education and living with children was associated with higher odds of increasing MVPA activity.

### RQ 4: Associations with changes in MSA

Being employed during lockdown and being overweight (BMI 25-29.99) or obese (BMI ≥30) was associated with higher odds of decreasing MSA activity. Older age (35-64, and 65+) and deterioration in psychological wellbeing were associated with lower odds of increasing MSA. See Table 5 for details.

**Table 5.** Research question 4: Independent associations of change in MSA. Segmented analyses of active sample (pre-Covid-19 MSA activity ≥1 day/week; predicting decrease by ≥1 day/week) and less inactive sample (pre-Covid-19 MSA activity 0 days/week; predicting increase by ≥1 day/week). Findings from fully adjusted logistic regression models on weighted data and using BH FDR adjustment (significant in bold).

| | Sample active ($\geq 1$ day/week)<br>pre-Covid-19 (weighted n=985) | | | | Sample inactive (0 days/week)<br>pre-Covid-19 (weighted n=1456) | | | |
| --- | --- | --- | --- | --- | --- | --- | --- | --- |
|  | Decrease (n=444) vs not |  |  |  | Increase (n=239) vs not |  |  |  |
|  | aOR | 95% CI |  | p | aOR | 95% CI |  | p |
| Female | 0.94 | 0.71 | 1.23 | 0.634 | 0.96 | 0.70 | 1.31 | 0.792 |
| Age (in decades) | 1.03 | 0.94 | 1.14 | 0.516 | - | - | - | - |
| Age $\leq 34$ | | | | | | | | |
| Age: 35-64 | - | - | - | - | **0.22** | **0.15** | **0.33** | **<0.001** |
| Age: 65+ | - | - | - | - | **0.34** | **0.20** | **0.58** | **<0.001** |
| White ethnicity | 0.75 | 0.51 | 1.11 | 0.154 | 0.71 | 0.41 | 1.23 | 0.216 |
| Household income $\geq 50,000$ | | | | | | | | |
| Income: $<50,000$ GBP | 1.11 | 0.80 | 1.55 | 0.535 | 0.67 | 0.45 | 0.99 | 0.047 |
| Income: Prefer not to say | 0.94 | 0.54 | 1.62 | 0.818 | 0.56 | 0.29 | 1.08 | 0.084 |
| Education: Highschool or higher | 0.82 | 0.59 | 1.15 | 0.260 | 1.20 | 0.85 | 1.70 | 0.310 |
| Employed | **1.81** | **1.34** | **2.46** | **<0.001** | 1.08 | 0.75 | 1.55 | 0.671 |
| Laid-off/Furloughed | 1.34 | 0.85 | 2.12 | 0.205 | 1.30 | 0.80 | 2.10 | 0.289 |
| Condition limiting PA | 1.11 | 0.72 | 1.72 | 0.632 | 0.64 | 0.38 | 1.05 | 0.078 |
| BMI $\leq 24.99$ | | | | | | | | |
| BMI: 25-29.99 | **1.88** | **1.39** | **2.55** | **<0.001** | 0.89 | 0.63 | 1.26 | 0.512 |
| BMI: Obese: 30+ | **3.38** | **2.23** | **5.14** | **<0.001** | 0.64 | 0.42 | 0.97 | 0.037 |
| Total isolation | 1.87 | 1.02 | 3.42 | 0.042 | 0.80 | 0.42 | 1.52 | 0.499 |
| Minor/no Covid-19 risk percept | 1.43 | 1.06 | 1.92 | 0.019 | 0.93 | 0.65 | 1.31 | 0.669 |
| Living with children | 1.01 | 0.71 | 1.44 | 0.964 | 1.48 | 0.94 | 2.31 | 0.088 |
| Living with vulnerable | 1.17 | 0.79 | 1.74 | 0.437 | 0.95 | 0.63 | 1.44 | 0.809 |
| Access garden/balcony to exercise comfortably in | 1.02 | 0.74 | 1.41 | 0.891 | 0.84 | 0.60 | 1.17 | 0.301 |
| Access green space within walking distance | 1.07 | 0.79 | 1.43 | 0.667 | 0.87 | 0.64 | 1.18 | 0.370 |
| Smoker | 0.63 | 0.43 | 0.91 | 0.013 | 1.04 | 0.74 | 1.46 | 0.828 |
| Weekly alcohol drinking, none (ref) |  |  |  |  |  |  |  |  |
| Weekly or less | 1.11 | 0.74 | 1.65 | 0.620 | 1.13 | 0.73 | 1.76 | 0.580 |
| More than weekly | 1.26 | 0.86 | 1.85 | 0.237 | 1.06 | 0.71 | 1.59 | 0.786 |
| Deteriorated psychological wellbeing | 1.27 | 0.97 | 1.66 | 0.082 | **0.62** | **0.46** | **0.83** | **0.002** |

Notes: All models were also adjusted for the time of enrolment and time when Covid-19 started to affect individuals in any way. Following the BH correction the highest p-value that met the threshold for significance for was $p=0.019$ .

## Discussion

This study shows that the first Covid-19 lockdown in the UK affected differently the MVPA and MSA levels, with different factors associated with engagement in and changes in these two activity types.

### Meeting WHO recommendations for MVPA and MSA post-Covid-19

Adherence to WHO recommendations for both MVPA and MSA pre- and post-Covid-19 was low – at about 15%. Older adults (aged 65+), adults who were obese, those with pre-Covid-19 household income below 50.000 GBP and who had conditions limiting physical activity were at risk of not meeting the WHO recommendations for both MVPA and MSA. These groups should be target by future interventions.

Moreover, meeting the recommendations for MVPA and MSA before Covid-19 was strongly predictive of meeting them post-Covid-19. Additionally, adults who remained active tended to maintain at least similar levels and forms of the exercises as pre-pandemic. Thus, as UK adults remain relatively consistent in their behaviour even during the lockdown, this study highlights the need to especially support those with low activity levels to develop healthy exercising routines.

### Changes to MVPA and MSA levels from pre- to the post-Covid-19 pandemic start

Although the group level data suggest little change in physical activity pre- to post-Covid-19, especially MSA, about a third of adults decreased and another third increased their MVPA from pre- to post-Covid-19. Those who changed MVPA decreased or increased their MVPA levels by over three and two hours per week, respectively. Those who changed MSA either gained or lost two days of MSA per week.

Decreases in MVPA was found among other adults samples in the UK^15^, Italy ^19^ and Spain^20^. However, the findings that over 70% of UK adults maintained or increased their MVPA could be at least partially attributed to the UK government’s consistent lockdown policies that allowed leaving the house for exercising, which placed UK in contrast to other countries^20,27.^

The maintenance of MSA levels could be at least partially explained by the low pre-Covid-19 MSA levels. Additionally, many MSA exercises can be performed with no equipment and at one’s home (e.g. Pilates, push-ups) and thus may be less affected by social distancing measures. This is further supported by the present findings that total isolation (i.e. not leaving the house for any reason) was associated with MVPA but not MSA levels.

### Factors associated with changes and maintenance of MVPA and MSA post-Covid-19

Older adults were more likely to maintain their pre-Covid-19 MVPA and MSA levels, including inactivity. Due to more established routines and lower baseline physical activity this group might have been less affected by the lockdown restrictions, such as gym closures or team sports bans^28^.

MVPA inactivity was more commonly maintained by those who were employed or living in total isolation. Such adults likely had fewer opportunities or flexibility to exercise. Additionally, those who had a health condition limiting physical activity and those living in total isolation tended to decrease MVPA. Factors that were associated with maintaining MVPA activity post-Covid-19 were white ethnicity, higher education, and having a garden or balcony large enough to exercise.

Increasing MVPA levels among those who were previously inactive was most common among those with higher education, which is in line with prior studies ^29^, and those who lived with children. Caring for children might have promoted some forms of MVPA, including outdoors, particularly as parents became responsible for children’s physical education during school lockdown closures.

In terms of changes in MSA levels, older adults were at higher risk of maintaining MSA inactivity, while adults with higher BMI levels were at greater risk of decreasing MSA from pre- to post-Covid-19. Therefore, both older adults and those with higher BMI should be among priority groups targeted with MSA interventions, particularly as MSA can bring important clinical benefits to these two groups^30-33^. Other priority groups for MSA training are those with lower household income and lower educational attainment.

Finally, as found previously^18^, psychological wellbeing was associated with physical activity levels. In the present study deterioration in psychological wellbeing was associated with not meeting guidelines for MVPA or decreasing MVPA levels, but not with MSA. Due to the cross-sectional design causality cannot be assumed, as poorer psychological wellbeing (e.g. lower mood, anxiety, stress), can be both a predictor and a consequence of low exercise levels^34,35.^ However, interventions aimed at improving physical activity are likely to improve mental health as well^34-36^.

### Strength and limitation

The study is among a few that assessed the levels and changes to both MVPA and MSA during the first UK lockdown. The study drew on previously used measures of MVPA and MSA^37^, which were supplemented by images to clarify exercise types. This study also benefits from a large list of correlates and covariates being measured to control for confounding. A number of sensitivity analyses were conducted to test the robustness of findings. The key limitations are that this was a cross-sectional study among self-selected sample that relied on self-report and recall that are prone to bias. Finally, the unfolding of the Covid-19 pandemic has coincided with season change from winter to spring. Without a true baseline from the same period in 2019 it is not possible to tease apart the effect of weather change from that of the pandemic.

## Conclusions

The findings point to social inequalities in how the first lockdown in the UK has affected physical activity levels, with differential impact on aerobic and strength training. Dedicated interventions are needed to support MVPA and MSA especially among those who have low activity levels, those who are older, have lower income, and have higher body mass index.

## Data Availability

Interested authors are encouraged to contact HEBECO co-leads at UCL to access data and to collaborate on future outputs: Dr Aleksandra Herbec and Prof Lion Shahab.

https://osf.io/sbgru/

https://osf.io/spdtb

https://osf.io/w7vbf/

https://osf.io/2ujxq/

## Footnotes

### Contributors

AH conceived the idea in consultation with PL, AB and LS. AH and VS analysed the data. All authors interpreted the data. AH drafted the first version of the manuscript. All authors critically revised the manuscript and approved the final draft before submission.

### Patient and public involvement

Patients and/or the public were not involved in the design, or conduct, or reporting, or dissemination plans of this research.

### Patient consent for publication

Obtained.

### Ethics approval

The study was approved by the by UCL Research Ethics Committee at the UCL Division of Psychology and Language Sciences (PaLS) (CEHP/2020/579; as part of the larger programme of work: The optimisation and implementation of interventions to change behaviours related to health and the environment).

## Acknowledgements

We are grateful to all participants who have been supporting our research. We would like to thank Public Health England, and particularly members of the Behavioural Insights at Public Health England for providing feedback on the survey wording. We would like to thank Public Health England, Cancer Research UK, local authorities, Mayors’ offices, as well as charities and other organisations in the UK, including the Asthma UK and British Lung Foundation Partnership, for supporting our recruitment campaign.

