## Supplementary Materials 1-5 for "Correlates of and changes in aerobic physical activity and strength training before and after the onset of COVID-19 pandemic in the UK – findings from the HEBECO study"

### Abstract

**Objectives:** Understanding changes in moderate to vigorous aerobic physical activity (MVPA) and strength training (MSA) from before to after (pre-/post-) the onset of the Covid-19 pandemic in the UK and their correlates can inform interventions.

**Methods:** We analysed retrospective and concurrent data on MVPA/MSA pre- and post-Covid-19 (up until 14th June 2020) among 2,657 UK adults. The associations between socio-demographic and health characteristics, MVPA/MSA pre-Covid-19, living and exercise conditions and meeting WHO recommended levels for MVPA/MSA/both (vs meeting neither), and changes in MVPA/MSA from pre- to post-Covid-19 were evaluated following stratification for pre-Covid-19 MVPA and MSA levels.

**Results:** A third of adults maintained (30.4%), decreased (36.2%) or increased (33.4%) their MVPA levels post-Covid-19. For MSA, the percentages were 61.6%, 18.2%, and 20.2%, respectively. MVPA increased or decreased by an average of 150min/week, and MSA by 2 days/week. Meeting both MSA+MVPA recommendations during lockdown (vs. meeting neither) was positively associated with meeting MVPA+MSA pre-lockdown (aOR=16.11,95%CI=11.24-23.07), and post-16-years of age education (aOR=1.57,1.14-2.17), and negatively associated with being obese (aOR=0.49,0.33-0.73), older age (65+ vs ≤34; aOR=.53,.32-.87), and annual household income <50.000GBP (vs ≥50.000GBP; aOR=0.65,0.46-0.91). The odds for decreasing MVPA were significantly lower for white ethnicity, post-16-years of age education, access to garden/balcony, and higher for those who were in total isolation. The odds for decreasing MSA were significantly higher for those who were overweight or obese.

**Conclusions:** Aerobic and strength training were differently impacted during the first UK lockdown, with poorer outcomes for older participants, those in isolation, and those with higher BMI.

**Keywords:** aerobic activity, strength training, covid-19, UK, cross-sectional, changes, MSA, MVPA

**Pre-registration:** https://osf.io/spdtb

### Supplementary Materials – HEBECO PA1 paper

### Supplementary Materials 1: Changes to the pre-registered protocol

The analysis protocol pre-registered on OSF prior to data inspection outlined analyses for research questions 3 and 4 involving unstratified multinomial analyses with change in MVPA and MSA as dependent variables. Specifically, the pre-registered protocol specified that we will use multinomial regression models to assess the correlates of a three-level dependent variables on change in (i) MVPA (increase by at least 20min/week, maintenance with change between 0 to 19min, and a decrease in MVPA by at least 20min/week) and in (ii) MSA (increase by at least 1 day/week, maintenance with change=0 days/week, and a decrease with change by at least 1 day/week). All the analyses were performed as per the original plan and can be accessed here: https://osf.io/2ujxq/.

Upon inspecting the data and the results a decision was made that this approach is not satisfactory as the group of maintainers of MVPA and MSA category included both inactive and active participants. This has motivated us to conduct and report the present analyses that stratified the sample based on pre-Covid-19 activity levels to identify meaningful changes.

### Supplementary Materials 2: detailed description of study measures

Below are the details of the all the explanatory measures assessed at baseline that are relevant for the study. For the full wording of the baseline survey see: https://osf.io/bja7g/

#### Socio-demographic, health and wellbeing characteristics

**Gender** (female gender/other; prefer not to say (PNS) were treated as missing) was assessed by asking participants: “Which of the following best describes how you think of yourself?”.

**Ethnicity** (white/non-white; PNS treated as missing) was assessed by asking participants: “What is your ethnic group?”.

**Education** (high school education or higher; yes/no) was assessed by asking participants: “What is the highest level of education that you have completed?”

**Employment** (employed or self-employed; yes/no) and **furloughed/laid off** (yes/no) were assessed by asking participants: “What is your current MAIN occupation (during COVID-19)?”.

**Income** (low-middle <50 000 GBP vs high ≥50 000 GBP vs PNS) was assessed by asking participants: “What was your household annual income BEFORE COVID-19? (do not include income of friends)”.

**Age** was assessed as a continuous variable. For the analyses we used age in decades and entered it as a continuous variable. Where the continuous variable violated the linearity assumptions of the logistic regression models (as did its logarithmic and square root transformation), a 3-level categorical variable was created (≤34, 35-64, 65+).

#### Health behaviours and conditions

- **Body mass index, BMI** (normal and underweight ≤24.99/overweight 25-29.99/ obese ≥30/ unknown (included answers ‘don’t know’ and PNS to height or weight variables)). BMI was calculated by dividing participants weight in kilogram by their squared height in meters and categorised into: normal weight and underweight. BMI is associated with both MVPA and MSA levels (Churilla et al., 2018).

- **Self-reports of any condition that limited their ability to engage in physical activity** (yes/no; PNS treated as missing). Participants were asked: “Do you now suffer from any condition that limits you from engaging in physical activity, including walking or doing housework?”

- **Self-perceived risk of Covid-19 to one’s health** (minor or no risk/major, significant, moderate risk or ‘don’t know’) was assessed with the question: “What risk does COVID-19 pose to your health?”

- **Smoking status** (current smokers vs not) was assessed using a single item: “Which statement about tobacco use and cigarette smoking best describes you?”. Those responding 1 (“I smoke cigarettes (including hand-rolled) every day”) and 2 (“I smoke cigarettes (including hand-rolled), but not every day”) were classified as current smokers. Smoking is associated with physical activity levels (Kaczynski, Manske, Mannell, & Grewal, 2008).

- **Weekly alcohol consumption** since the Covid-19 pandemic has started was assessed by asking participants how many times they had a drink containing alcohol, with the categorical answers converted to weekly frequencies from 0 to 6.5 times per week (0=I have not had any alcohol after COVID-19 started, 0.25=monthly or less, 0.75=2 to 4 times a month, 2.5=2 to 3 times a week, 4.5=4 to 5 times a week, 6.5=6 or more times a week). In models where the continuous alcohol frequency variable violated the linearity assumptions of the logistic regression models (as did its logarithmic and square root transformation), weekly alcohol frequency was categorised into three levels (none/weekly or less often/more frequently than weekly). Alcohol drinking is associated with physical activity levels (Leasure, Neighbors, Henderson, & Young, 2015).

- **Deterioration in psychological wellbeing** (yes/no). Deterioration in psychological wellbeing was assessed on a five-point Likert scale (1=poor, 3=average, 5=excellent) by asking participants: "How would you rate the below aspects of your life? – Psychological wellbeing". The question was asked about the period before Covid-19 and since Covid-19. A change score was calculated by deducting the score before from since score. As the continuous variable violated the linearity assumptions of the logistic regression models (as did its logarithmic and square root transformation), participants were categorised into either deteriorated psychological wellbeing (change score $< 0$) versus maintained or improved psychological wellbeing (change score $\geq 0$).

- **Isolation status** (total isolation/other) was assessed by asking which type of Covid-19 induced isolation participants were experiencing. Answers were dichotomised into indications of total isolation/quarantine ("not leaving the house for any reasons, not even to buy groceries or medications or to exercise") versus any other (some, general, or no isolation). Participants' perceived risk of Covid-19 to their health was dichotomised into 'no or minor risk perception' versus any other indication (moderate risk, significant, major or not known). It was hypothesised that perceived risk could affect physical activity, especially MVPA that may happen outdoors.

- **Access to space to exercise** was assessed using a single question on participants' access to spaces that allowed multiple answers, including for: „a garden or balcony big enough to exercise comfortably" and to "a public park/green space that is within a walking distance and open during Covid-19“. The answers were dummy coded into separate binary variables (yes/no).

- **Living with children age 15 and younger** (yes/no) and **living with vulnerable persons** (any of: persons over the age of 70, persons in poor health versus, or persons who they believed may be vulnerable to Covid-19 for any reason) was assessed by asking participants: "Do you live with any of these persons below? (select all that apply)." It was hypothesised that living with these groups could limit some of the opportunities to exercise.

### Supplementary materials 3: Univariable analyses.

**Supplementary Table S3.1.** (univariable) Research question 2 -Predictors of meeting both, MVPA only and MSA only in comparison to meeting WHO guidelines for neither MVPA nor MSA (reference, n=1178). Results from unadjusted models on weighted data with BH FDR correction of p-values (significant in bold).

| | Both (n=388) | | | | MVPA only (n=570) | | | | MSA only (n=306) | | | |
|---|---|---|---|---|---|---|---|---|---|---|---|---|
| | OR | 95% CI | | *p* | OR | 95% CI | | *p* | OR | 95% CI | | *p* |
| Female | 1.01 | 0.80 | 1.27 | 0.928 | **0.62** | **0.51** | **0.76** | **<0.001** | 0.93 | 0.72 | 1.19 | 0.546 |
| Age ≤34[a] | | | | | | | | | | | | |
| Age: 35-64 | **0.57** | **0.43** | **0.75** | **<0.001** | 1.08 | 0.82 | 1.42 | 0.604 | **0.41** | **0.30** | **0.56** | **<0.001** |
| Age: 65+ | **0.41** | **0.28** | **0.60** | **<0.001** | 1.02 | 0.73 | 1.43 | 0.893 | **0.58** | **0.40** | **0.84** | **0.004** |
| White ethnicity | 0.77 | 0.52 | 1.12 | 0.166 | 1.53 | 1.02 | 2.30 | 0.040 | **0.43** | **0.30** | **0.62** | **<0.001** |
| Household income ≥50.000 | | | | | | | | | | | | |
| Income: <50 000 GBP | **0.44** | **0.33** | **0.59** | **<0.001** | **0.66** | **0.50** | **0.85** | **0.002** | 0.80 | 0.57 | 1.12 | 0.194 |
| Income: Prefer not to say | **0.41** | **0.25** | **0.66** | **<0.001** | **0.50** | **0.32** | **0.77** | **0.002** | 0.79 | 0.47 | 1.34 | 0.385 |
| Education: Highschool or higher | **2.57** | **1.96** | **3.38** | **<0.001** | **1.34** | **1.09** | **1.65** | **0.006** | **2.41** | **1.79** | **3.24** | **<0.001** |
| Employed | 1.14 | 0.90 | 1.43 | 0.272 | 1.20 | 0.98 | 1.46 | 0.077 | 1.05 | 0.81 | 1.35 | 0.714 |
| Laid-off/Furloughed | 0.95 | 0.67 | 1.35 | 0.784 | 1.01 | 0.75 | 1.36 | 0.972 | 1.04 | 0.72 | 1.51 | 0.839 |
| Condition limiting PA | **0.24** | **0.16** | **0.37** | **<0.001** | **0.27** | **0.19** | **0.37** | **<0.001** | **0.64** | **0.46** | **0.89** | **0.009** |
| BMI ≤24 | | | | | | | | | | | | |
| BMI: 25-29.99 | **0.62** | **0.48** | **0.81** | **<0.001** | 0.86 | 0.68 | 1.08 | 0.191 | **0.60** | **0.45** | **0.79** | **<0.001** |
| BMI: Obese: 30+ | **0.31** | **0.22** | **0.43** | **<0.001** | **0.62** | **0.48** | **0.81** | **<0.001** | **0.27** | **0.19** | **0.40** | **<0.001** |
| Total isolation | **0.30** | **0.17** | **0.52** | **<0.001** | **0.17** | **0.10** | **0.31** | **<0.001** | 0.67 | 0.43 | 1.05 | 0.079 |
| Minor/no Covid-19 risk percept | **1.70** | **1.34** | **2.16** | **<0.001** | **1.39** | **1.12** | **1.72** | **0.002** | **1.37** | **1.05** | **1.79** | **0.019** |
| Living with children | 0.99 | 0.73 | 1.33 | 0.925 | 0.81 | 0.61 | 1.06 | 0.121 | 0.70 | 0.49 | 1.01 | 0.056 |
| Living with vulnerable | 1.09 | 0.80 | 1.48 | 0.583 | 0.80 | 0.59 | 1.06 | 0.122 | 0.87 | 0.61 | 1.25 | 0.450 |
| Access garden/balcony to exercise comfortably in | 1.18 | 0.91 | 1.54 | 0.212 | 1.01 | 0.81 | 1.27 | 0.918 | 0.82 | 0.63 | 1.08 | 0.157 |
| Access green space within walking distance | **1.70** | **1.33** | **2.16** | **<0.001** | **1.59** | **1.29** | **1.95** | **<0.001** | 1.29 | 1.00 | 1.67 | 0.049 |
| Smoker | **0.48** | **0.35** | **0.64** | **<0.001** | **0.72** | **0.57** | **0.91** | **0.006** | **0.64** | **0.47** | **0.87** | **0.004** |
| Alcohol drinking frequency, never (ref) | | | | | | | | | | | | |
| Weekly or less | 1.32 | 0.94 | 1.85 | 0.106 | **1.50** | **1.12** | **2.02** | **0.006** | 1.48 | 1.04 | 2.11 | 0.032 |
| More than weekly | **1.47** | **1.09** | **1.98** | **0.012** | **1.50** | **1.15** | **1.95** | **0.003** | 1.21 | 0.87 | 1.68 | 0.249 |
| Deteriorated psychological wellbeing | **0.62** | **0.50** | **0.79** | **<0.001** | **0.71** | **0.58** | **0.87** | **0.001** | 0.74 | 0.58 | 0.95 | 0.019 |
| Meeting WHO PA rec (meeting neither=ref) | | | | | | | | | | | | |
| Meeting both before | **17.58** | **12.53** | **24.65** | **<0.001** | **2.30** | **1.60** | **3.31** | **<0.001** | **4.03** | **2.72** | **5.99** | **<0.001** |
| Meeting only MVPA before | **4.53** | **3.13** | **6.55** | **<0.001** | **8.30** | **6.47** | **10.65** | **<0.001** | 1.67 | 1.08 | 2.58 | 0.021 |
| Meeting only MSA before | **7.72** | **5.27** | **11.32** | **<0.001** | 0.86 | 0.52 | 1.42 | 0.549 | **10.31** | **7.37** | **14.43** | **<0.001** |

**Supplementary Table S3.2.** (univariable) Research question 3: Independent associations of change in MVPA. Segmented analyses of active sample (pre-Covid-19 MVPA activity ≥30min; predicting decrease by ≥20min) and less active/inactive sample (pre-Covid-19 MVPA activity <30min; predicting increase by ≥20min). Findings from unadjusted logistic regression models on weighted data and using BH FDR adjustment (significant in bold).

| | Sample active (≥30min/week) pre-Covid-19 (weighted n=1857) | | | | Sample inactive (<30min/week) pre-Covid-19 (weighted n=585) | | | |
|---|---|---|---|---|---|---|---|---|
| | Decrease (n=868) vs not | | | | Increase (n=174) vs not | | | |
| | OR | 95% CI | | *p* | OR | 95% CI | | *p* |
| Female | **1.38** | **1.15** | **1.65** | **0.001** | 1.13 | 0.79 | 1.62 | 0.496 |
| Age (in decades) | 0.94 | 0.88 | 0.99 | 0.024 | **0.67** | **0.59** | **0.75** | **<0.001** |
| White ethnicity | **0.57** | **0.42** | **0.77** | **<0.001** | 0.46 | 0.24 | 0.88 | 0.019 |
| Household income ≥50.000 | | | | | | | | |
| Income: <50 000 GBP | **1.36** | **1.08** | **1.71** | **0.010** | **0.45** | **0.27** | **0.77** | **0.003** |
| Income: Prefer not to say | **1.59** | **1.09** | **2.33** | **0.017** | 0.53 | 0.25 | 1.14 | 0.103 |
| Education: Highschool or higher | **0.72** | **0.59** | **0.88** | **0.001** | **2.67** | **1.81** | **3.93** | **<0.001** |
| Employed | 0.82 | 0.69 | 0.99 | 0.036 | 0.80 | 0.55 | 1.16 | 0.235 |
| Laid-off/Furloughed | 1.22 | 0.92 | 1.62 | 0.165 | **2.16** | **1.36** | **3.43** | **0.001** |
| Condition limiting PA | **2.28** | **1.71** | **3.04** | **<0.001** | **0.27** | **0.16** | **0.43** | **<0.001** |
| BMI ≤24 | | | | | | | | |
| BMI: 25-29.99 | 1.01 | 0.82 | 1.23 | 0.961 | 1.04 | 0.69 | 1.55 | 0.863 |
| BMI: Obese: 30+ | 0.99 | 0.78 | 1.27 | 0.945 | 0.56 | 0.34 | 0.90 | 0.016 |
| Total isolation | **4.95** | **3.11** | **7.88** | **<0.001** | **0.22** | **0.10** | **0.49** | **<0.001** |
| Minor/no Covid-19 risk percept | 0.94 | 0.77 | 1.14 | 0.503 | **2.17** | **1.50** | **3.14** | **<0.001** |
| Living with children | 0.89 | 0.70 | 1.13 | 0.319 | 1.37 | 0.82 | 2.30 | 0.225 |
| Living with vulnerable | **0.72** | **0.56** | **0.93** | **0.012** | 1.14 | 0.70 | 1.85 | 0.599 |
| Access garden/balcony to exercise comfortably in | 0.82 | 0.67 | 1.00 | 0.053 | **0.61** | **0.42** | **0.89** | **0.011** |
| Access green space within walking distance | 0.81 | 0.67 | 0.98 | 0.026 | **1.66** | **1.16** | **2.37** | **0.006** |
| Smoker | 0.94 | 0.75 | 1.17 | 0.576 | 1.15 | 0.79 | 1.68 | 0.454 |
| Alcohol drinking frequency, never (ref) | | | | | | | | |
| Weekly or less | **0.68** | **0.52** | **0.89** | **0.005** | 1.19 | 0.73 | 1.96 | 0.483 |
| More than weekly | **0.60** | **0.47** | **0.76** | **<0.001** | 1.22 | 0.78 | 1.90 | 0.387 |
| Deteriorated psychological wellbeing | **1.42** | **1.18** | **1.71** | **<0.001** | 1.03 | 0.73 | 1.47 | 0.854 |

Adjusted for the time of enrolment and time when Covid-19 started to affect individuals in any way.

**Supplementary Table S3.3** Research question 4: Independent associations of change in MSA. Segmented analyses of active sample (pre-Covid-19 MSA activity ≥1 day/week; predicting decrease by ≥1 day/week) and less inactive sample (pre-Covid-19 MSA activity 0 days/week; predicting increase by ≥1 day/week). Findings from unadjusted logistic regression models on weighted data and using BH FDR adjustment (significant in bold).

| | Sample active (≥1 day/week) pre-Covid-19 (weighted n=985) | | | | Sample inactive (0 days/week) pre-Covid-19 (weighted n=1456) | | | |
|---|---|---|---|---|---|---|---|---|
| | Decrease (n=444) vs not | | | | Increase (n=239) vs not | | | |
| | OR | 95% CI | | *p* | OR | 95% CI | | *p* |
| Female | 0.87 | 0.67 | 1.12 | 0.274 | 0.85 | 0.64 | 1.12 | 0.249 |
| Age (in decades) | 1.02 | 0.94 | 1.10 | 0.625 | - | - | - | - |
| Age ≤34[a] | | | | | | | | |
| Age: 35-64 | - | - | - | - | **0.20** | **0.14** | **0.28** | **<0.001** |
| Age: 65+ | - | - | - | - | **0.27** | **0.18** | **0.40** | **<0.001** |
| White ethnicity | 0.80 | 0.57 | 1.13 | 0.210 | **0.36** | **0.22** | **0.58** | **<0.001** |
| Household income ≥50.000 | | | | | | | | |
| Income: <50 000 GBP | 1.01 | 0.75 | 1.36 | 0.953 | **0.61** | **0.42** | **0.87** | **0.007** |
| Income: Prefer not to say | 0.80 | 0.48 | 1.31 | 0.368 | 0.64 | 0.36 | 1.17 | 0.147 |
| Education: Highschool or higher | 0.74 | 0.55 | 1.00 | 0.051 | **1.86** | **1.37** | **2.52** | **<0.001** |
| Employed | **1.60** | **1.24** | **2.06** | **<0.001** | 1.01 | 0.76 | 1.33 | 0.951 |
| Laid-off/Furloughed | 0.99 | 0.67 | 1.45 | 0.954 | 1.33 | 0.91 | 1.96 | 0.141 |
| Condition limiting PA | 1.35 | 0.92 | 1.97 | 0.120 | **0.43** | **0.27** | **0.67** | **<0.001** |
| BMI ≤24 | | | | | | | | |
| BMI: 25-29.99 | **1.88** | **1.42** | **2.50** | **<0.001** | 0.68 | 0.50 | 0.93 | 0.016 |
| BMI: Obese: 30+ | **3.20** | **2.18** | **4.72** | **<0.001** | **0.43** | **0.29** | **0.63** | **<0.001** |
| Total isolation | **2.05** | **1.22** | **3.45** | **0.007** | 0.62 | 0.35 | 1.10 | 0.102 |
| Minor/no Covid-19 risk percept | 1.07 | 0.83 | 1.38 | 0.596 | **1.61** | **1.20** | **2.15** | **0.001** |
| Living with children | 0.83 | 0.60 | 1.15 | 0.266 | 1.44 | 0.95 | 2.18 | 0.086 |
| Living with vulnerable | 1.13 | 0.79 | 1.63 | 0.498 | 1.01 | 0.69 | 1.47 | 0.971 |
| Access garden/balcony to exercise comfortably in | 0.90 | 0.67 | 1.19 | 0.453 | 0.74 | 0.55 | 0.99 | 0.043 |
| Access green space within walking distance | 0.94 | 0.72 | 1.22 | 0.621 | 1.12 | 0.85 | 1.49 | 0.420 |
| Smoker | 0.65 | 0.46 | 0.90 | 0.010 | 1.00 | 0.74 | 1.36 | 0.995 |
| Alcohol drinking frequency, never (ref) | | | | | | | | |
| Weekly or less | 0.88 | 0.61 | 1.27 | 0.508 | 1.50 | 1.00 | 2.27 | 0.051 |
| More than weekly | 0.93 | 0.67 | 1.30 | 0.677 | 1.40 | 0.97 | 2.03 | 0.072 |
| Deteriorated psychological wellbeing | 1.37 | 1.06 | 1.77 | 0.014 | 0.72 | 0.55 | 0.95 | 0.021 |

Adjusted for the time of enrolment and time when Covid-19 started to affect individuals in any way.

[a]Age was categorised in models where the continuous predictor violated the linearity assumption.

### Supplementary Materials 4: Unweighted analyses

**Supplementary Table S4.1.** (unweighted data) Sample characteristics % (N).

| | Included sample (2657) | Excluded sample (336) | *p* |
|---|---|---|---|
| Female | 68.9 (1831) | 68.1 (224) | 0.760 |
| Age; M (SD) | 48.4 (15.2) | 43.7 (16.7) | <0.001 |
| White ethnicity | 94.4 (2507) | 91.1 (297) | 0.020 |
| Household income ≥50.000 | 90.9 (993) | 9.1 (100) | <0.001 |
| Income: <50 000 GBP | 54.5 (1447) | 55.5 (186) | |
| Income: Prefer not to say | 8.2 (217) | 14.6 (49) | |
| Education: Highschool or higher | 86.8 (2307) | 85.7 (288) | 0.571 |
| Employed | 56.1 (1490) | 49.1 (164) | 0.016 |
| Laid-off/Furloughed | 9.8 (261) | 12.3 (41) | 0.161 |
| Condition limiting PA | 13.2 (351) | 19.6 (54) | 0.003 |
| BMI ≤24 | 49.1 (1305) | 23.8 (62) | <0.001 |
| BMI: 25-29.99 | 31.3 (832) | 15.8 (41) | |
| BMI: Obese: 30+ | 19.6 (520) | 9.2 (24) | |
| Total isolation | 7.2 (190) | 7.2 (21) | 0.955 |
| Minor/no Covid-19 risk percept | 37.5 (996) | 42.4 (123) | 0.101 |
| Living with children | 19.6 (520) | 19.0 (64) | 0.820 |
| Living with vulnerable | 14.8 (394) | 16.1 (54) | 0.547 |
| Access garden/balcony | 71.9 (1911) | 63.1 (212) | 0.001 |
| Access green space | 63.5 (1687) | 57.1 (192) | 0.023 |
| Time affected up till mid March | 49.5 (1316) | 55.1 (185) | 0.056 |
| Enrolled from 1st June (ref) | 5.2 (139) | 4.8 (16) | 0.479 |
| Enrolled up until 15th May | 48.9 (1298) | 45.8 (154) | |
| Enrolled second half of May | 45.9 (1220) | 49.4 (166) | |
| Smoker | 17.7 (470) | 25.6 (86) | <0.001 |
| Alcohol drinking frequency, never (ref) | 17.0 (453) | 19.6 (66) | 0.092 |
| Weekly or less | 27.5 (732) | 31.3 (105) | |
| More than weekly | 55.4 (1472) | 49.1 (165) | |
| Deteriorated psychological wellbeing | 55.1 (1465) | 50.2 (117) | 0.148 |
| Meeting WHO PA rec (meeting both=ref) | 17.9 (475) | 14.9 (26) | 0.332 |
| Meeting only MVPA before | 22.8 (605) | 23.0 (40) | |
| Meeting only MSA before | 14.9 (395) | 11.5 (20) | |
| Meeting neither before | 44.5 (1182) | 50.6 (88) | |

**Supplementary Table S4.2.** (unweighted data) Aerobic physical activity (MVPA) and strength training (MSA) before to after the Covid-19 pandemic has started to affect individuals in the UK (up until 14th June 2020).

| | Before Covid-19[1] | Since Covid-19[1] |
|---|---|---|
| Inactive (0 sessions MVPA and 0 days MSA) % (N) | 14.1 (375) | 16.0 (425) |
| MVPA 0 sessions % (N) | 15.9 (422) | 18.5 (491) |
| MVPA | | |
| MVPA min/week among active; Median (IQR) | 135.0 (180.0) | 150.0 (240.0) |
| Mean (SD) | 215.3 (363.4) | 227.0 (341.7) |
| Mean (SD) replacing extremes with 840 min/week | 191.8 (186.9) | 206.9 (193.7) |
| MVPA min/week among entire sample; Median (IQR) | 120.0 (185.0) | 120.0 (215.0) |
| Mean (SD) | 184.9 (345.0) | 190.7 (324.1) |
| Mean (SD) replacing extremes with 840 min/week | 164.7 (185.6) | 173.8 (193.1) |
| MSA Median (IQR) | 0 (2) | 0 (2) |
| MSA % (N) | | |
| 0 das/week | 53.0 (1408) | 54.0 (1435) |
| 1 day/week | 14.3 (379) | 11.7 (311) |
| 2 days/week | 14.6 (387) | 10.5 (280) |
| 3 days/week | 10.4 (277) | 9.6 (254) |
| 4+ days/week | 7.8 (206) | 14.2 (377) |
| Meet guidelines % (N) | | |
| Neither MVPA nor MSA | 44.5 (1182) | 42.0 (1117) |
| Meet MVPA (≥150min/week) only | 22.8 (605) | 23.7 (629) |
| Meet MSA (≥2 days/week) only | 14.9 (395) | 14.7 (390) |
| Meets both MVPA and MSA | 17.9 (475) | 19.6 (521) |
| Change in form of exercise % (N) | | |
| None of the same | | 13.2 (350) |
| Some of the same | | 29.1 (774) |
| About half the same | | 8.8 (233) |
| Mostly the same | | 23.4 (622) |
| Exactly the same | | 10.0 (267) |

**Supplementary Table S4.3.** (unweighted data) Research question 2 – Factors associated with meeting both, MVPA only and MSA only in comparison to meeting WHO guidelines for neither MVPA nor MSA (reference, n=1117) after the Covid-19 pandemic has started to affect individuals in the UK (up until June 2020[1]). Results from fully adjusted models on unweighted data with BH FDR correction of p-values (significant in bold).

| | Both (n=521) | | | | MVPA only (n=629) | | | | MSA only (n=390) | | | |
|---|---|---|---|---|---|---|---|---|---|---|---|---|
| Characteristics and associations | aOR | 95% CI | | *p* | aOR | 95% CI | | *p* | aOR | 95% CI | | *p* |
| Female | 1.17 | 0.89 | 1.52 | 0.256 | 0.93 | 0.73 | 1.18 | 0.546 | 1.16 | 0.87 | 1.53 | 0.308 |
| Age ≤34[a] | | | | | | | | | | | | |
| Age: 35-64 | 0.92 | 0.67 | 1.25 | 0.589 | **1.49** | **1.09** | **2.05** | **0.013** | **0.59** | **0.43** | **0.82** | **0.001** |
| Age: 65+ | 0.64 | 0.40 | 1.03 | 0.067 | **1.80** | **1.17** | **2.75** | **0.007** | 0.90 | 0.57 | 1.44 | 0.664 |
| White ethnicity | 1.36 | 0.81 | 2.27 | 0.245 | 1.79 | 1.01 | 3.15 | 0.044 | 0.67 | 0.42 | 1.08 | 0.099 |
| Household income ≥50.000 | | | | | | | | | | | | |
| Income: <50 000 GBP | **0.58** | **0.44** | **0.76** | **<0.001** | **0.70** | **0.55** | **0.90** | **0.006** | 1.00 | 0.75 | 1.33 | 0.976 |
| Income: Prefer not to say | **0.55** | **0.34** | **0.89** | **0.014** | 0.76 | 0.49 | 1.17 | 0.214 | 0.90 | 0.54 | 1.48 | 0.667 |
| Education: Highschool or higher | 1.38 | 0.92 | 2.09 | 0.121 | 1.20 | 0.86 | 1.67 | 0.293 | **1.86** | **1.19** | **2.90** | **0.007** |
| Employed | 0.78 | 0.58 | 1.05 | 0.102 | 1.11 | 0.85 | 1.47 | 0.442 | 1.22 | 0.89 | 1.68 | 0.212 |
| Laid-off/Furloughed | 1.00 | 0.64 | 1.55 | 0.997 | 1.07 | 0.70 | 1.62 | 0.759 | 1.33 | 0.84 | 2.13 | 0.227 |
| Condition limiting PA | **0.42** | **0.27** | **0.65** | **<0.001** | **0.36** | **0.24** | **0.54** | **<0.001** | 0.78 | 0.53 | 1.16 | 0.219 |
| BMI ≤24 | | | | | | | | | | | | |
| BMI: 25-29.99 | **0.61** | **0.46** | **0.80** | **<0.001** | 0.80 | 0.62 | 1.02 | 0.076 | **0.66** | **0.49** | **0.88** | **0.005** |
| BMI: Obese: 30+ | **0.35** | **0.24** | **0.51** | **<0.001** | **0.69** | **0.51** | **0.93** | **0.014** | **0.43** | **0.30** | **0.62** | **<0.001** |
| Total isolation | **0.39** | **0.22** | **0.69** | **0.001** | **0.22** | **0.12** | **0.41** | **<0.001** | 0.84 | 0.52 | 1.38 | 0.498 |
| Minor/no Covid-19 risk percept | 0.82 | 0.64 | 1.07 | 0.147 | 0.92 | 0.72 | 1.17 | 0.476 | 0.90 | 0.68 | 1.20 | 0.477 |
| Living with children | 0.79 | 0.58 | 1.07 | 0.130 | **0.56** | **0.42** | **0.76** | **<0.001** | 0.89 | 0.64 | 1.24 | 0.497 |
| Living with vulnerable | 1.14 | 0.81 | 1.60 | 0.457 | 1.07 | 0.79 | 1.46 | 0.662 | 0.94 | 0.65 | 1.35 | 0.721 |
| Access garden/balcony to exercise comfortably in | 1.09 | 0.82 | 1.44 | 0.562 | 1.00 | 0.78 | 1.29 | 0.994 | 0.98 | 0.74 | 1.31 | 0.901 |
| Access green space within walking distance | 1.17 | 0.90 | 1.51 | 0.243 | 1.23 | 0.97 | 1.56 | 0.081 | 1.16 | 0.89 | 1.53 | 0.278 |
| Smoker | **0.56** | **0.40** | **0.79** | **0.001** | **0.69** | **0.52** | **0.93** | **0.016** | **0.65** | **0.46** | **0.92** | **0.016** |
| Alcohol drinking frequency, never (ref) | | | | | | | | | | | | |
| Weekly or less | 1.24 | 0.84 | 1.82 | 0.279 | 1.16 | 0.82 | 1.63 | 0.405 | 1.07 | 0.72 | 1.58 | 0.745 |
| More than weekly | 1.25 | 0.88 | 1.78 | 0.221 | 1.14 | 0.83 | 1.56 | 0.412 | 1.02 | 0.71 | 1.47 | 0.923 |
| Deteriorated psychological wellbeing | **0.61** | **0.48** | **0.78** | **<0.001** | 0.78 | 0.63 | 0.98 | 0.029 | **0.65** | **0.51** | **0.84** | **0.001** |
| Meeting WHO PA rec (meeting neither=ref) | | | | | | | | | | | | |
| Meeting both before | **14.58** | **10.48** | **20.30** | **<0.001** | **2.47** | **1.74** | **3.52** | **<0.001** | **3.53** | **2.44** | **5.10** | **<0.001** |
| Meeting only MVPA before | **3.31** | **2.35** | **4.66** | **<0.001** | **5.86** | **4.54** | **7.55** | **<0.001** | 0.84 | 0.54 | 1.31 | 0.445 |
| Meeting only MSA before | **4.98** | **3.49** | **7.11** | **<0.001** | 0.79 | 0.51 | 1.24 | 0.303 | **6.59** | **4.80** | **9.06** | **<0.001** |

Models adjusted for the time of enrolment and time when Covid-19 started to affect individuals in any way.

[1] Covering the period of the strictest first lockdown in the UK

**Supplementary Table S4.4.** Research question 3: Independent associations of change in MVPA from before to after the Covid-19 pandemic has started to affect individuals in the UK (up until 14th June 2020[2]). Segmented analyses of active sample (pre-Covid-19 MVPA activity ≥30min; predicting decrease by ≥20min) and less active/inactive sample (pre-Covid-19 MVPA activity <30min; predicting increase by ≥20min). Findings from fully adjusted logistic regression models on unweighted data and using BH FDR adjustment (significant in bold).

| Characteristics and associations | Sample active (≥30min/week) pre-Covid-19 (unweighted n=2132) | | | | Sample inactive (<30min/week) pre-Covid-19 (unweighted n=525) | | | |
|---|---|---|---|---|---|---|---|---|
| | Decrease (n=905) vs not | | | | Increase (n=195) vs not | | | |
| | aOR | 95% CI | | *p* | aOR | 95% CI | | *p* |
| Female | 1.05 | 0.86 | 1.27 | 0.660 | 1.12 | 0.73 | 1.74 | 0.605 |
| Age (in decades) | **0.91** | **0.85** | **0.98** | **0.011** | **0.80** | **0.68** | **0.94** | **0.007** |
| White ethnicity | **0.52** | **0.35** | **0.76** | **0.001** | 0.80 | 0.30 | 2.15 | 0.656 |
| Household income ≥50.000 | | | | | | | | |
| Income: <50 000 GBP | **1.41** | **1.15** | **1.73** | **0.001** | 0.95 | 0.59 | 1.53 | 0.829 |
| Income: Prefer not to say | 1.31 | 0.92 | 1.87 | 0.130 | 0.72 | 0.32 | 1.65 | 0.441 |
| Education: Highschool or higher | 0.73 | 0.54 | 0.98 | 0.036 | 1.87 | 1.07 | 3.26 | 0.028 |
| Employed | 0.96 | 0.77 | 1.18 | 0.682 | 0.70 | 0.43 | 1.13 | 0.140 |
| Laid-off/Furloughed | 0.95 | 0.68 | 1.34 | 0.782 | 1.51 | 0.77 | 2.93 | 0.227 |
| Condition limiting PA | **1.87** | **1.37** | **2.55** | **<0.001** | **0.47** | **0.27** | **0.83** | **0.010** |
| BMI ≤24 | | | | | | | | |
| BMI: 25-29.99 | 1.08 | 0.88 | 1.33 | 0.456 | 1.11 | 0.69 | 1.79 | 0.665 |
| BMI: Obese: 30+ | 0.97 | 0.75 | 1.26 | 0.833 | 0.71 | 0.42 | 1.19 | 0.195 |
| Total isolation | **2.91** | **1.89** | **4.46** | **<0.001** | **0.22** | **0.09** | **0.54** | **0.001** |
| Minor/no Covid-19 risk percept | 1.21 | 0.99 | 1.47 | 0.067 | 1.19 | 0.76 | 1.87 | 0.441 |
| Living with children | 0.80 | 0.64 | 1.01 | 0.061 | 0.86 | 0.52 | 1.44 | 0.570 |
| Living with vulnerable | 0.92 | 0.71 | 1.20 | 0.551 | 0.83 | 0.49 | 1.43 | 0.511 |
| Access garden/balcony to exercise comfortably in | **0.74** | **0.60** | **0.91** | **0.005** | 0.96 | 0.61 | 1.49 | 0.847 |
| Access green space within walking distance | 0.95 | 0.78 | 1.15 | 0.589 | 1.45 | 0.96 | 2.19 | 0.077 |
| Smoker | 0.88 | 0.68 | 1.13 | 0.311 | 1.70 | 1.05 | 2.74 | 0.030 |
| Alcohol drinking frequency, never (ref) | | | | | | | | |
| Weekly or less | 1.08 | 0.81 | 1.44 | 0.605 | 0.96 | 0.54 | 1.71 | 0.890 |
| More than weekly | 0.89 | 0.68 | 1.16 | 0.378 | 1.24 | 0.75 | 2.06 | 0.399 |
| Deteriorated psychological wellbeing | **1.40** | **1.16** | **1.68** | **<0.001** | 1.12 | 0.74 | 1.67 | 0.597 |

Models adjusted for the time of enrolment and time when Covid-19 started to affect individuals in any way.

[2] Covering the period of the strictest first lockdown in the UK

**Supplementary Table S4.5.** (unweighted data). Research question 4: Independent associations of change in MSA from before to after the Covid-19 pandemic has started to affect individuals in the UK (up until 14th June 2020[3]). Segmented analyses of active sample (pre-Covid-19 MSA activity ≥1 day/week; predicting decrease by ≥1 day/week) and less inactive sample (pre-Covid-19 MSA activity 0 days/week; predicting increase by ≥1 day/week). Findings from fully adjusted logistic regression models on unweighted data and using BH FDR adjustment (significant in bold).

| | Sample active (≥1 day/week) pre-Covid-19 (unweighted n=1249) | | | | Sample inactive (0 days/week) pre-Covid-19 (unweighted n=1408) | | | |
|---|---|---|---|---|---|---|---|---|
| | Decrease (n=534) vs not | | | | Increase (n=284) vs not | | | |
| Characteristics and associations | aOR | 95% CI | | *p* | aOR | 95% CI | | *p* |
| Female | 0.83 | 0.64 | 1.08 | 0.160 | 1.216 | 0.899 | 1.647 | 0.205 |
| Age (in decades) | 1.03 | 0.94 | 1.12 | 0.579 | - | - | - | - |
| Age ≤34[a] | | | | | | | | |
| Age: 35-64 | - | - | - | - | **0.353** | **0.248** | **0.502** | **<0.001** |
| Age: 65+ | - | - | - | - | **0.305** | **0.178** | **0.524** | **<0.001** |
| White ethnicity | 0.89 | 0.57 | 1.39 | 0.611 | 0.553 | 0.298 | 1.024 | 0.060 |
| Household income ≥50.000 | | | | | | | | |
| Income: <50 000 GBP | 1.25 | 0.96 | 1.64 | 0.096 | **0.678** | **0.496** | **0.928** | **0.015** |
| Income: Prefer not to say | 1.39 | 0.88 | 2.18 | 0.155 | 0.480 | 0.258 | 0.893 | 0.020 |
| Education: Highschool or higher | 0.96 | 0.62 | 1.49 | 0.863 | 1.310 | 0.837 | 2.051 | 0.237 |
| Employed | **1.49** | **1.12** | **1.97** | **0.005** | 0.900 | 0.634 | 1.278 | 0.557 |
| Laid-off/Furloughed | 1.18 | 0.75 | 1.85 | 0.467 | 1.104 | 0.676 | 1.803 | 0.694 |
| Condition limiting PA | 1.08 | 0.72 | 1.62 | 0.708 | 0.664 | 0.409 | 1.078 | 0.097 |
| BMI ≤24 | | | | | | | | |
| BMI: 25-29.99 | **1.63** | **1.24** | **2.13** | **<0.001** | 0.846 | 0.613 | 1.168 | 0.310 |
| BMI: Obese: 30+ | **2.50** | **1.73** | **3.60** | **<0.001** | 0.707 | 0.484 | 1.031 | 0.072 |
| Total isolation | **2.03** | **1.18** | **3.50** | **0.011** | 0.861 | 0.481 | 1.543 | 0.615 |
| Minor/no Covid-19 risk percept | 1.18 | 0.91 | 1.52 | 0.216 | 1.045 | 0.769 | 1.421 | 0.777 |
| Living with children | 1.04 | 0.77 | 1.41 | 0.784 | 0.708 | 0.503 | 0.997 | 0.048 |
| Living with vulnerable | 1.11 | 0.78 | 1.59 | 0.553 | 0.990 | 0.677 | 1.446 | 0.957 |
| Access garden/balcony to exercise comfortably in | 0.86 | 0.65 | 1.13 | 0.275 | 0.765 | 0.562 | 1.040 | 0.088 |
| Access green space within walking distance | 0.95 | 0.74 | 1.23 | 0.713 | 1.102 | 0.816 | 1.489 | 0.527 |
| Smoker | **0.61** | **0.43** | **0.87** | **0.007** | 1.096 | 0.772 | 1.557 | 0.608 |
| Alcohol drinking frequency, never (ref) | | | | | | | | |
| Weekly or less | 1.01 | 0.69 | 1.49 | 0.943 | 1.192 | 0.776 | 1.833 | 0.422 |
| More than weekly | 0.99 | 0.69 | 1.42 | 0.951 | 1.124 | 0.755 | 1.672 | 0.565 |
| Deteriorated psychological wellbeing | **1.38** | **1.09** | **1.76** | **0.008** | **0.643** | **0.488** | **0.848** | **0.002** |

Models adjusted for the time of enrolment and time when Covid-19 started to affect individuals in any way.
[a]Age was categorised in models where the continuous predictor violated the linearity assumption.

[3] Covering the period of the strictest first lockdown in the UK

**Supplementary Materials 5: Sensitivity Analyses for RQ3 (different cut-off values for MVPA change).**

**Supplementary Table S5.1.** Predicting MVPA decrease of at least 15 min in the active sample from before to after the Covid-19 pandemic has started to affect individuals in the UK (up until 14th June 2020). (fully adjusted model, weighted data).

| Characteristics and associations | **aOR** | **95% CI** | | ***p*** |
|---|---|---|---|---|
| Female | 1.232 | 1.010 | 1.503 | .039 |
| Age (in decades) | **.914** | **.849** | **.983** | **.015** |
| White ethnicity | **.649** | **.464** | **.909** | **.012** |
| Household income ≥50.000 | 1.0 | | | |
| Income: <50 000 GBP | 1.272 | .986 | 1.641 | .064 |
| Income: Prefer not to say | 1.182 | .783 | 1.783 | .427 |
| Education: Highschool or higher | **.754** | **.602** | **.944** | **.014** |
| Employed | .841 | .677 | 1.045 | .118 |
| Laid-off/Furloughed | 1.025 | .738 | 1.423 | .884 |
| Condition limiting PA | **1.732** | **1.260** | **2.381** | **.001** |
| BMI ≤24 | 1.0 | | | |
| BMI: 25-29.99 | 1.025 | .822 | 1.280 | .824 |
| BMI: Obese: 30+ | .875 | .667 | 1.147 | .334 |
| Total isolation | **3.682** | **2.252** | **6.020** | **.000** |
| Minor/no Covid-19 risk percept | .984 | .791 | 1.224 | .882 |
| Living with children | .858 | .661 | 1.114 | .251 |
| Living with vulnerable | .801 | .607 | 1.058 | .118 |
| Access garden/balcony to exercise comfortably in | .773 | .618 | .968 | .025 |
| Access green space within walking distance | .813 | .662 | .998 | .048 |
| Smoker | 1.106 | .861 | 1.420 | .432 |
| Alcohol drinking frequency, never (ref) | 1.0 | | | |
| Weekly or less | .790 | .596 | 1.049 | .103 |
| More than weekly | .772 | .594 | 1.003 | .053 |
| Deteriorated psychological wellbeing | **1.422** | **1.170** | **1.728** | **.000** |

Model was adjusted for all the variable sin the table as well as the time of enrolment and time when Covid-19 started to affect individuals in any way. Significant results are in bold.

**Supplementary Table S5.2.** Predicting MVPA increase of at least 15 min in the active sample from before to after the Covid-19 pandemic has started to affect individuals in the UK (up until 14th June 2020; fully adjusted model, weighted data).

| Characteristics and associations | **aOR** | **95% CI** | | **p** |
|---|---|---|---|---|
| Female | 1.513 | 1.003 | 2.282 | .048 |
| Age (in decades) | **.750** | **.642** | **.876** | **.000** |
| White ethnicity | .564 | .249 | 1.276 | .169 |
| Household income ≥50.000 | 1.0 | | | |
| Income: <50 000 GBP | .633 | .339 | 1.182 | .152 |
| Income: Prefer not to say | .663 | .271 | 1.620 | .367 |
| Education: Highschool or higher | 1.700 | 1.078 | 2.683 | .023 |
| Employed | .846 | .526 | 1.362 | .492 |
| Laid-off/Furloughed | 1.380 | .762 | 2.498 | .288 |
| Condition limiting PA | **.445** | **.249** | **.795** | **.006** |
| BMI ≤24 | 1.0 | | | |
| BMI: 25-29.99 | 1.640 | 1.014 | 2.652 | .044 |
| BMI: Obese: 30+ | .778 | .451 | 1.343 | .368 |
| Total isolation | .395 | .172 | .907 | .029 |
| Minor/no Covid-19 risk percept | 1.349 | .857 | 2.122 | .196 |
| Living with children | 1.600 | .917 | 2.793 | .098 |
| Living with vulnerable | .872 | .505 | 1.508 | .624 |
| Access garden/balcony to exercise comfortably in | .730 | .469 | 1.136 | .163 |
| Access green space within walking distance | .986 | .656 | 1.481 | .944 |
| Smoker | 1.491 | .963 | 2.309 | .073 |
| Alcohol drinking frequency, never (ref) | 1.0 | | | |
| Weekly or less | .719 | .407 | 1.272 | .257 |
| More than weekly | .941 | .567 | 1.563 | .814 |
| Deteriorated psychological wellbeing | 1.275 | .850 | 1.913 | .241 |

Model was adjusted for all the variable sin the table as well as the time of enrolment and time when Covid-19 started to affect individuals in any way. Significant results are in bold.

**Supplementary Table S5.3**. Predicting MVPA decrease of at least 30 min in the active sample (fully adjusted model, weighted data).

| Characteristics and associations | **aOR** | **95% CI** | | **p** |
|---|---|---|---|---|
| Female | 1.207 | .989 | 1.474 | .064 |
| Age (in decades) | **.904** | **.841** | **.973** | **.007** |
| White ethnicity | **.618** | **.441** | **.865** | **.005** |
| Household income ≥50.000 | 1.0 | | | |
| Income: <50 000 GBP | 1.356 | 1.049 | 1.753 | .020 |
| Income: Prefer not to say | 1.276 | .845 | 1.927 | .246 |
| Education: Highschool or higher | **.741** | **.592** | **.928** | **.009** |
| Employed | .853 | .686 | 1.061 | .153 |
| Laid-off/Furloughed | 1.037 | .747 | 1.441 | .827 |
| Condition limiting PA | **1.672** | **1.219** | **2.293** | **.001** |
| BMI ≤24 | 1.0 | | | |
| BMI: 25-29.99 | 1.047 | .838 | 1.308 | .684 |
| BMI: Obese: 30+ | .880 | .671 | 1.155 | .357 |
| Total isolation | **3.426** | **2.130** | **5.511** | **.000** |
| Minor/no Covid-19 risk percept | .978 | .785 | 1.218 | .842 |
| Living with children | .837 | .644 | 1.087 | .182 |
| Living with vulnerable | .756 | .572 | .998 | .048 |
| Access garden/balcony to exercise comfortably in | **.744** | **.594** | **.932** | **.010** |
| Access green space within walking distance | .820 | .667 | 1.008 | .059 |
| Smoker | 1.097 | .854 | 1.410 | .468 |
| Alcohol drinking frequency, never (ref) | 1.0 | | | |
| Weekly or less | .753 | .567 | .999 | .049 |
| More than weekly | .752 | .578 | .977 | .033 |
| Deteriorated psychological wellbeing | **1.403** | **1.154** | **1.706** | **.001** |

Model was adjusted for all the variable sin the table as well as the time of enrolment and time when Covid-19 started to affect individuals in any way. Significant results are in bold.

**Supplementary Table S5.4.** Predicting MVPA increase of at least 30 min in the inactive sample (fully adjusted model, weighted data).

| Characteristics and associations | **aOR** | **95% CI** | | **p** |
|---|---|---|---|---|
| Female | **1.877** | **1.213** | **2.902** | **.005** |
| Age (in decades) | **.707** | **.599** | **.836** | **.000** |
| White ethnicity | 1.391 | .575 | 3.363 | .464 |
| Household income ≥50.000 | 1.0 | | | |
| Income: <50 000 GBP | **.394** | **.205** | **.757** | **.005** |
| Income: Prefer not to say | **.208** | **.077** | **.560** | **.002** |
| Education: Highschool or higher | **2.116** | **1.296** | **3.456** | **.003** |
| Employed | **.516** | **.309** | **.861** | **.011** |
| Laid-off/Furloughed | 1.380 | .757 | 2.517 | .293 |
| Condition limiting PA | .478 | .257 | .887 | .019 |
| BMI ≤24 | 1.0 | | | |
| BMI: 25-29.99 | 1.511 | .917 | 2.490 | .105 |
| BMI: Obese: 30+ | .616 | .341 | 1.113 | .108 |
| Total isolation | .332 | .133 | .827 | .018 |
| Minor/no Covid-19 risk percept | 1.001 | .620 | 1.616 | .997 |
| Living with children | **2.290** | **1.225** | **4.283** | **.009** |
| Living with vulnerable | .936 | .519 | 1.687 | .825 |
| Access garden/balcony to exercise comfortably in | .764 | .482 | 1.209 | .250 |
| Access green space within walking distance | .960 | .622 | 1.481 | .853 |
| Smoker | .979 | .622 | 1.542 | .926 |
| Alcohol drinking frequency, never (ref) | 1.0 | | | |
| Weekly or less | 1.028 | .555 | 1.904 | .931 |
| More than weekly | 1.234 | .708 | 2.152 | .459 |
| Deteriorated psychological wellbeing | .888 | .579 | 1.362 | .585 |

Model was adjusted for all the variable sin the table as well as the time of enrolment and time when Covid-19 started to affect individuals in any way. Significant results are in bold.
